# APOE4 Copy Number-Dependent Proteomic changes in the Cerebrospinal Fluid

**DOI:** 10.1101/2020.06.30.20143578

**Authors:** Miles Berger, Mary Cooter, Alexander S. Roesler, Stacey Chung, John Park, Jennifer L. Modliszeski, Keith W. VanDusen, J. Will Thompson, Arthur Moseley, Michael J. Devinney, Shayan Smani, Ashley Hall, Victor Cai, Jeffrey N. Browndyke, Michael W. Lutz, David L. Corcoran, Alzheimer’s Disease Neuroimaging Initiative

## Abstract

**Background:** *APOE4* has been hypothesized to increase Alzheimer’s disease risk by increasing neuroinflammation, though the specific neuroinflammatory pathways involved are unclear.

**Objectives:** To characterize CSF proteomic changes as a function of *APOE4* copy number.

**Methods:** We analyzed targeted proteomic data obtained on ADNI CSF samples using a linear regression model adjusting for age, sex, and *APOE4* copy number, and a second linear model also adjusting for AD clinical status. False Discovery Rate (FDR) was used to correct for multiple comparisons.

**Results:** In the first model, increasing *APOE4* copy number was associated with significant expression decreases in a CRP peptide (q=0.006), and significant expression increases in peptides from ALDOA, CH3L1 (YKL-40), and FABPH (q<0.05 for each). In the second model (controlling for age, sex, and AD clinical status), increasing *APOE4* copy number was associated with significant expression decreases in a CRP peptide (q=0.009). In both models, increased *APOE4* copy number was associated with trends towards lower expression of all 24 peptides from all 8 different complement proteins measured here, although none of these differences were statistically significant. The odds of this happening by chance for 24 unrelated peptides would be less than 1 in 16 million.

**Conclusions:** Increasing *APOE4* copy number was associated with decreased CSF CRP levels and increased CSF ALDOA, CH3L1 and FABH levels; the CRP decrease remained significant after controlling for AD clinical status. Increased *APOE4* copy number may also be associated with decreased CSF complement pathway protein levels, a hypothesis for investigation in future studies.

## Introduction

The best described genetic contributor to late onset Alzheimer’s disease (LOAD) is the e4 polymorphism of the apolipoprotein E gene [1]. Individuals carrying a single *APOE4* allele copy have a ∼3-fold increased risk of developing Alzheimer’s disease (AD), and those who carry two *APOE4* alleles have a greater than 10-fold risk of developing AD [2-5]. Additionally, the presence of an *APOE4* allele is associated with worse neurologic outcomes including a higher index of disability in multiple sclerosis patients, worse cognitive outcomes following mild traumatic brain injury, and increased risk of death following subarachnoid hemorrhage [6-8], as well as increased atherosclerotic cardiovascular disease risk [9]. Likely due to these pleiotropic effects, *APOE4* carriers live ∼4.2 years less than non-*APOE4* carriers [10, 11]. Despite our knowledge of these multiple negative effects of *APOE4*, it remains unclear what the mechanisms are that explain how *APOE4* contributes to AD risk and worse outcomes across these other disease states.

The APOE protein has multiple biological roles, including cholesterol transport in the central nervous system (CNS), signaling through cell surface receptors, and modulating synaptic function by regulating the expression of syntaxin-1, PSD95, and NMDA and AMPA receptors [12]. Given this multitude of functions, it is unclear which mechanisms explain the increased AD risk in *APOE4* carriers. Patients with an *APOE4* allele are typically diagnosed with AD in their 7^th^ or 8^th^ decade of life, even though the APOE protein is expressed within the CNS throughout life [13]. This suggests that *APOE4* likely contributes to AD risk before cognitive deficits first appear [14, 15]. This idea is supported by fMRI studies demonstrating that young adult *APOE4* carriers without AD have significant alterations in the default mode network when compared to non-carrier controls [16]. Additionally, young adult *APOE4* carriers show increased activation of the bilateral medial temporal lobe during an encoding task [17], which may be a compensatory mechanism to achieve normal cognitive function in *APOE4* carriers.

One mechanism hypothesized to underlie the link between *APOE4* and AD is neuroinflammation. Indeed, neuroinflammation is a key contributor to AD pathogenesis in humans [18]. Recent evidence in murine models has shown that human *APOE4* knock-in mice have increased glial activation in response to intra-cerebroventricular LPS injection and increased IL-1β, IL-6 and TNFα levels when compared to APOE2 or APOE3 allele knock-in mice [19]. Furthermore, microglia isolated from *APOE4/4* targeted replacement mice, as compared to those from *APOE3/3* mice, have increased pro-inflammatory cytokines such as IL-6, TNFα and IL12p40 [20, 21]. Although these studies have linked *APOE4* to increases in multiple inflammatory cytokines and pathways, it is unclear which of these inflammatory mechanisms are responsible for increased AD risk in *APOE4* carriers.

Because neurodegeneration in AD itself is associated with inflammation, it is important to study the effect of *APOE4* on the CNS of older adults who don’t yet have dementia and neurodegeneration due to AD. Such studies provide an opportunity to discover how *APOE4* affects the CNS and how it increases AD risk, before frank AD-related neuro-degeneration begins. Thus, here we analyzed targeted CSF proteomic data from Alzheimer’s Disease Neuroimaging Institute (ADNI) research subjects, while controlling for AD clinical status, in order to find CSF protein level variation associated with *APOE4* allele copy number.

## Methods

### ADNI study and participants

The patient data and clinical annotations used in this study were obtained from the Alzheimer’s Disease Neuroimaging Initiative (ADNI) database (adni.loni.usc.edu). ADNI is a longitudinal multicenter study that tracks and evaluates changes in cognition, brain structure and function, and biomarkers associated with the progression of mild cognitive impairment (MCI) and Alzheimer’s disease [22]. Further detail on ADNI is found in the Acknowledgments section. Each ADNI site received written informed consent from all participants and institutional review board approval.

Inclusion and exclusion criteria for the normal control (NC), MCI, and AD cohorts is available at adni.loni.usc.edu. Briefly, NC subjects were defined as having a mini-mental state examination (MMSE) [23] score ≥ 24 and Clinical Dementia Rating (CDR) [24] score of 0 and having no confounding neurological or psychological disorders. MCI subjects had MMSE scores of 23-30, a CDR score of 0.5, objective memory loss as measured by Wechsler Memory Scale Revised—Logical Memory II [25], and preserved activities of daily living. AD patients met the National Institute of Neurological and Communicative Disorders and Stroke and the Alzheimer’s Disease and Related Disorders Association (NINCDS-ADRDA) [26] criteria for probable AD and had MMSE scores of 20-26 and CDR scores of 0.5-1.0. The current study includes CSF samples from 289 unique ADNI-1 subjects (85 normal control, 134 MCI, and 66 AD patients). Publicly available metadata such as age, gender, diagnosis at baseline, MMSE score, and *APOE4* genotype were collected from the ADNI database. Cohort demographics are summarized in Table 1.

**Table 1:** Baseline characteristics of ADNI Patients, grouped by number of *APOE4* alleles Values represent means (SD), or percentages in the case of gender (for females), or count per group (for AT classification).

|  | **0 *APOE4* alleles**<br>**(N=148)** | **1 *APOE4* allele**<br>**(N=104)** | **2 *APOE4* alleles**<br>**(N=35)** | **p-value** |
| --- | --- | --- | --- | --- |
| **Age** | 75.94 (6.88) | 75.43 (6.77) | 71.86 (6.88) | 0.007<sup>1</sup> |
| **Gender (Male)** | 89 (60.1%) | 63 (60.6%) | 20 (57.1%) | 0.935<sup>2</sup> |
| **Race** |  |  |  | 0.473<sup>3</sup> |
| Asian | 3 (2.0%) | 0 (0.0%) | 0 (0.0%) |  |
| Black/African American | 5 (3.4%) | 5 (4.8%) | 0 (0.0%) |  |
| White | 140 (94.6%) | 99 (95.2%) | 35 (100.0%) |  |
| **Years of Education** | 16 [14, 18] | 16 [14, 18] | 16 [14, 16] | 0.227<sup>4</sup> |
| **CSF Aβ\*** | 988.47 (397.74) | 643.16 (208.95) | 482.48 (160.98) | <0.001<sup>1</sup> |
| **CSF Tau\*\*** | 273.08 (115.89) | 335.57 (109.98) | 348.32 (120.79) | <0.001<sup>1</sup> |
| **CSF p-tau\*\*** | 25.72 (12.59) | 33.81 (12.73) | 35.64 (15.21) | <0.001<sup>1</sup> |
| **Clinical Status** |  |  |  | <0.001<sup>2</sup> |
| Normal | 65 (43.9%) | 19 (18.3%) | 2 (5.7%) |  |
| MCI | 64 (43.2%) | 53 (51.0%) | 18 (51.4%) |  |
| AD | 19 (12.8%) | 32 (30.8%) | 15 (42.9%) |  |
| **ATN Classification\*\*\*<sup>+</sup>** |  |  |  | <0.001<sup>3</sup> |
| A-T- | 59 (40.4%) | 8 (7.8%) | 0 (0.0%) |  |
| A+T- | 37 (25.3%) | 26 (25.5%) | 9 (25.7%) |  |
| A-T+ | 13 (8.9%) | 3 (2.9 %) | 0 (0.0%) |  |
| A+T+ | 37 (25.3%) | 65 (63.7%) | 26 (74.3%) |  |

P-value key: 1= ANOVA, 2= Chi-square, 3=Fisher's Exact, 4=Kruskal Wallis
\*28 patients not included who returned values >1700; 4 patients with no BL CSF measures.
\*\* 4 patients with no BL CSF measures
<sup>+</sup> A+ defined as Aβ values below 1065 pg/ml, T+ defined as p-tau values over 27 pg/ml

### ADNI-1 CSF Collection and Processing

CSF samples (0.5 mL) were obtained at ADNI visits, stored, transported, and processed according to published procedures [27, 28]. Technical details on the mass spectrometry platform data acquisition, quality control metrics, and validation protocols used in this study are described in the ADNI “Use of Targeted Multiplex Proteomic Strategies to Identify Novel CSF Biomarkers in AD” data primer and in [29]. Briefly, CSF samples were depleted of high abundance proteins using MARS-14 immunoaffinity resin, trypsin digested (1:10 protease:protein ratio), lyophilized, and desalted prior to LC/MRM-MS proteomic analysis on a QTRAP 5500 LC-MS/MS system. CSF multiplex multiple reaction monitoring (MRM) is a standardized peptide panel developed as a QC metric to verify the reproducibility of sample processing and mass spectrometry analysis [30]. A total of 320 peptides produced by tryptic digestion of 143 proteins were identified and met the QC criteria of the ADNI working group for inclusion in the original dataset [29]. These peptides were selected to measure the levels of proteins previously implicated in AD neuropathology and/or neuro-inflammation [29].

Patients in the ADNI proteomics study were classified by AT (i.e. amyloid and p-tau) status, using previously reported CSF Aβ and p-tau measurements made with the Roche Elecsys platform, and previously described Aβ, and p-tau thresholds [31, 32]. We used the AT schema rather than the full ATN classification, because CSF tau and p-tau levels are highly co-linear, such that every patient who would be T+ would also be N+ and *vice versa* (Shaw LM, personal communication, 6/16/2020).

### Statistical analysis

Mass spectrometry data from the ADNI study was re-analyzed to compare peptide data by *APOE4* allele count. Peptides with an expression value below zero were set to missing values. The intraclass correlation (ICC) across technical replicates was calculated for each peptide. Subsequent analysis included 294 peptides that had an ICC >= 0.6. The technical replicate for each individual with the smallest number of missing peptides was used in the analysis. We analyzed CSF targeted proteomic data from 289 research participants in the ADNI-1 study, 85 of whom were healthy controls, 134 of whom had MCI, and 66 of whom had dementia due to AD. Association between each of the variables of interest with each peptide was tested in a linear model framework with an empirical Bayes method for parameter estimation from the limma [33] Bioconductor [34] package. Age and gender were included as cofactors in both models; AD clinical status (i.e. normal, MCI or dementia due to AD) was also included in model 2. False discovery rate was used to correct for multiple hypothesis testing.

## Results

### ADNI Patient Cohort Characteristics

Baseline characteristics of the ADNI-1 patients whose samples were used for targeted proteomics measurements [29] are presented in Table 1. Subjects with 0, 1 or 2 copies of the *APOE4* allele were similar in terms of gender, race and years of education. Consistent with prior work showing that the *APOE4* allele is associated with reduced longevity [11], individuals with two *APOE4* allele copies were ∼4 years younger those with zero or one copy of the *APOE4* allele. As expected, the percentage of patients with MCI and AD increased among patients with either 1 or 2 *APOE4* alleles. Consistent with prior work [35, 36], increasing *APOE4* copy number was associated with lower CSF Aβ levels. Increasing *APOE4* copy number was also associated with increases in CSF tau and p-tau levels (Table 1). Lastly, increasing *APOE4* copy number was associated with decreases in the proportion of patients who were A^-^T^-^ and increases in the proportion who were A^+^T^+^ (Table 1).

### Model 1: CSF proteomic changes and APOE4 gene dosage

To identify protein-derived peptides whose level(s) differ as a function of *APOE4* copy number, a linear model correcting for age and gender was used to test the relationship between *APOE4* copy number and CSF peptide levels. Initial analysis evaluated 294 peptides with sufficient replicability (ICC >= 0.6) for measuring CSF expression variance by *APOE4* copy number (Table 2). In this model, 12 of 294 peptides had significant expression changes (*q* <= 0.05) associated with increasing *APOE4* copy number (Table 3 and Figure 1A). CSF levels of an *APOE4*-specific peptide (APOE_LGADMEDVR) were substantially higher in *APOE4* carriers vs. non-carriers (*q* = 6.53 x 10^−81^), which is consistent with prior studies [37, 38]. Two peptides found in all APOE isotypes had elevated CSF expression with increasing *APOE4* copy number (APOE_LAVYQAGAR, *q* = 0.027; APOE_LGPLVEQGR, *q* = 0.027), whereas CSF expression of an APOE2-specific peptide was found to decrease with higher *APOE4* gene dosage (APOE_CLAVYQAGAR, *q* = 0.027). Increasing *APOE4* allele copy number was associated with reduced expression of a peptide from the acute inflammatory marker C-reactive protein (CRP) (*q* = 0.006). Increasing *APOE4* copy number was associated with increasing expression of peptides derived from the glycoprotein Chitinase 3-like protein 1 (CH3L1; also known as YKL-40) (*q* < 0.05), the cardiac injury biomarker heart-type fatty acid binding protein (FABPH) (*q* < 0.05) and the glycolytic enzyme fructose-bisphosphate aldolase A (ALDOA; *q* < 0.05).

**Table 2:** Proteins included in the ADNI Targeted CSF Proteomics Study

|  |  |  |  |  |
| --- | --- | --- | --- | --- |
| 1433Z | CMGA | IFNB | NELL2 | SCG3 |
| A1AT | CNDP1 | IGSF8 | NEO1 | SDCB1 |
| A1AT | CNTF | IL10 | NEUS | SE6L1 |
| A1BG | CNTN1 | IL12B | NFH | SHSA7 |
| A2GL | CNTN2 | IL17 | NFL | SIAE |
| A2MG | CO2 | IL1A | NFM | SLIK1 |
| A4 | CO3 | IL27A | NGF | SMOC1 |
| AACT | CO4A | IL6 | NICA | SODC |
| AATM | CO5 | IL6RA | NLGN3 | SODE |
| AFAM | CO6 | ITIH1 | NPTX1 | SORC1 |
| ALDOA | CO8B | ITIH5 | NPTX2 | SORC2 |
| AMBP | COCH | ITM2B | NPTXR | SORC3 |
| AMD | CRP | JAK1 | NPY | SPON1 |
| APLP2 | CSTN1 | KAIN | NRCAM | SPRL1 |
| APOA | CSTN3 | KCC2B | NRX1A | STX12 |
| APOA1 | CUTA | KI67 | NRX2A | SV2A |
| APOB | CYTC | KLK10 | NRX3A | SYNJ1 |
| APOC1 | DAG1 | KLK11 | NSG1 | SYT11 |
| APOD | DIAC | KLK12 | OSTP | TADBP |
| APOE | ENOG | KLK3 | PCD17 | TAU |
| B2MG | ENPP2 | KLK6 | PCMD1 | TCRG1 |
| B3GN1 | EXTL2 | KLK9 | PCSK1 | TEN3 |
| BACE1 | FABP5 | KLKB1 | PDIA3 | TGFB1 |
| BASP1 | FABP6 | KNG1 | PDYN | TGFB2 |
| BDNF | FABP7 | KPCZ | PEDF | TGFB3 |
| BTD | FABPH | KPYM | PGRP2 | TGON2 |
| C1QA | FABPI | L1CAM | PIMT | THRB |
| C1QB | FAM3C | LAMB2 | PLDX1 | TIMP1 |
| C3AR | FBLN1 | LFTY2 | PLMN | TNF14 |
| CA2D1 | FBLN3 | LPHN1 | PPN | TNFA |
| CAD13 | FETUA | LRC4B | PRDX1 | TNR1B |
| CADM3 | FMOD | LTBP2 | PRDX2 | TNR21 |
| CAH1 | GFAP | MIME | PRDX3 | TNR6 |
| CATA | GLNA | MMP2 | PRDX4 | TRBM |
| CATD | GOGB1 | MMP9 | PRDX5 | TRFE |
| CATL1 | GOLM1 | MMRN2 | PRDX6 | TRFM |
| CCKN | GRIA4 | MOG | PTGDS | TTHY |
| CCL25 | HBA | MTHR | PTPRD | UBB |
| CD14 | HBB | MUC18 | PTPRN | UCHL1 |
| CD59 | HEMO | NBL1 | PVRL1 | VASN |
| CERU | HERC4 | NCAM1 | RIMS3 | VGF |
| CFAB | I18BP | NCAM2 | SAP | VTDB |
| CH3L1 | IBP2 | NCAN | SCG1 | X3CL1 |
| CLUS | IBP6 | NEGR1 | SCG2 |  |

**Table 3:**
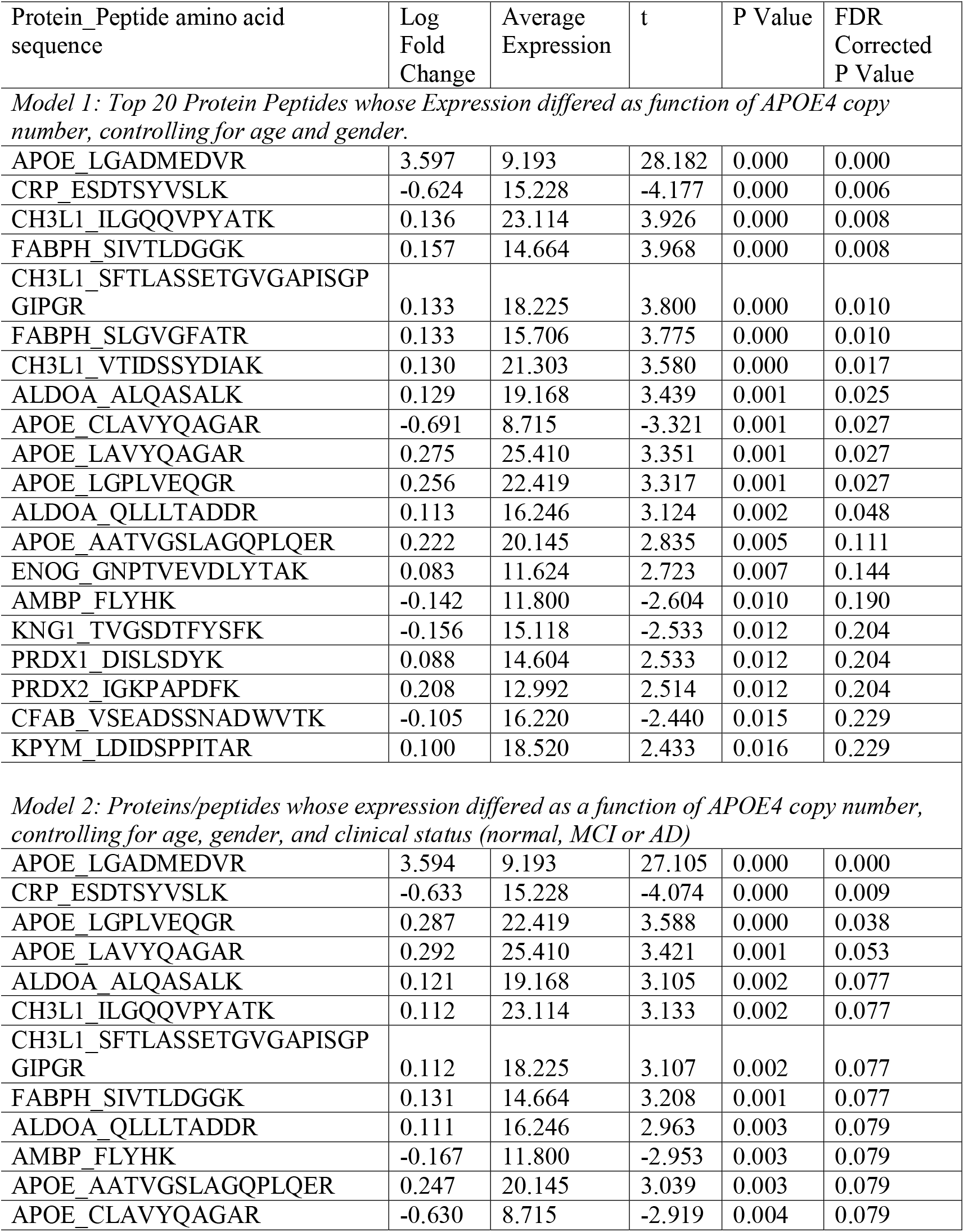
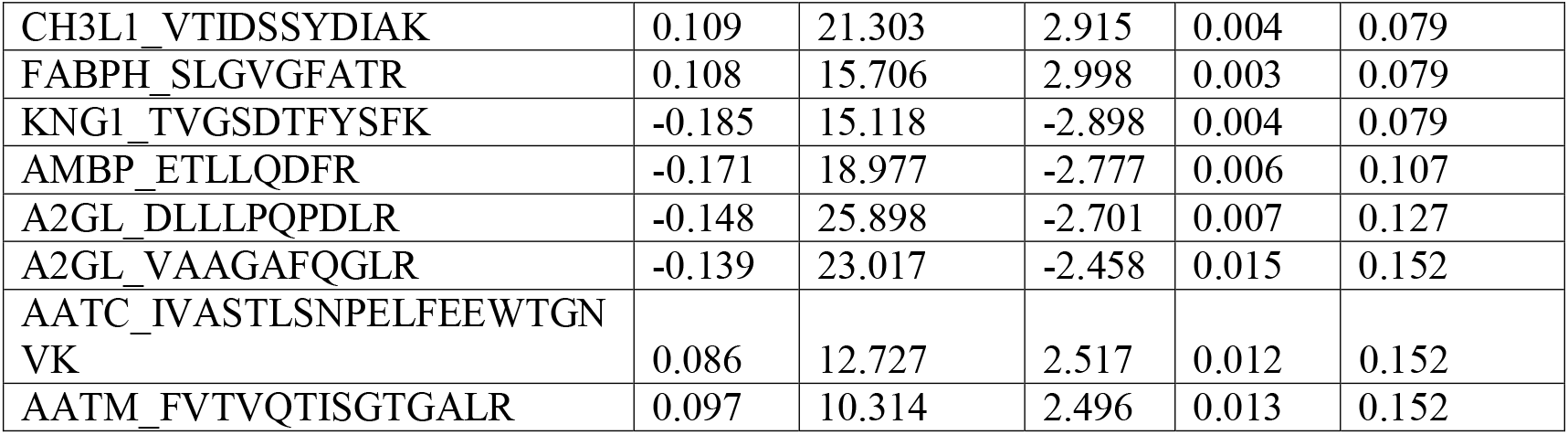
Summary of Expression for the Indicated Peptides/proteins by *APOE4* copy number in multivariate models accounting for age and gender (model 1) or age, gender and clinical status (Model 2).

**Fig 1.**
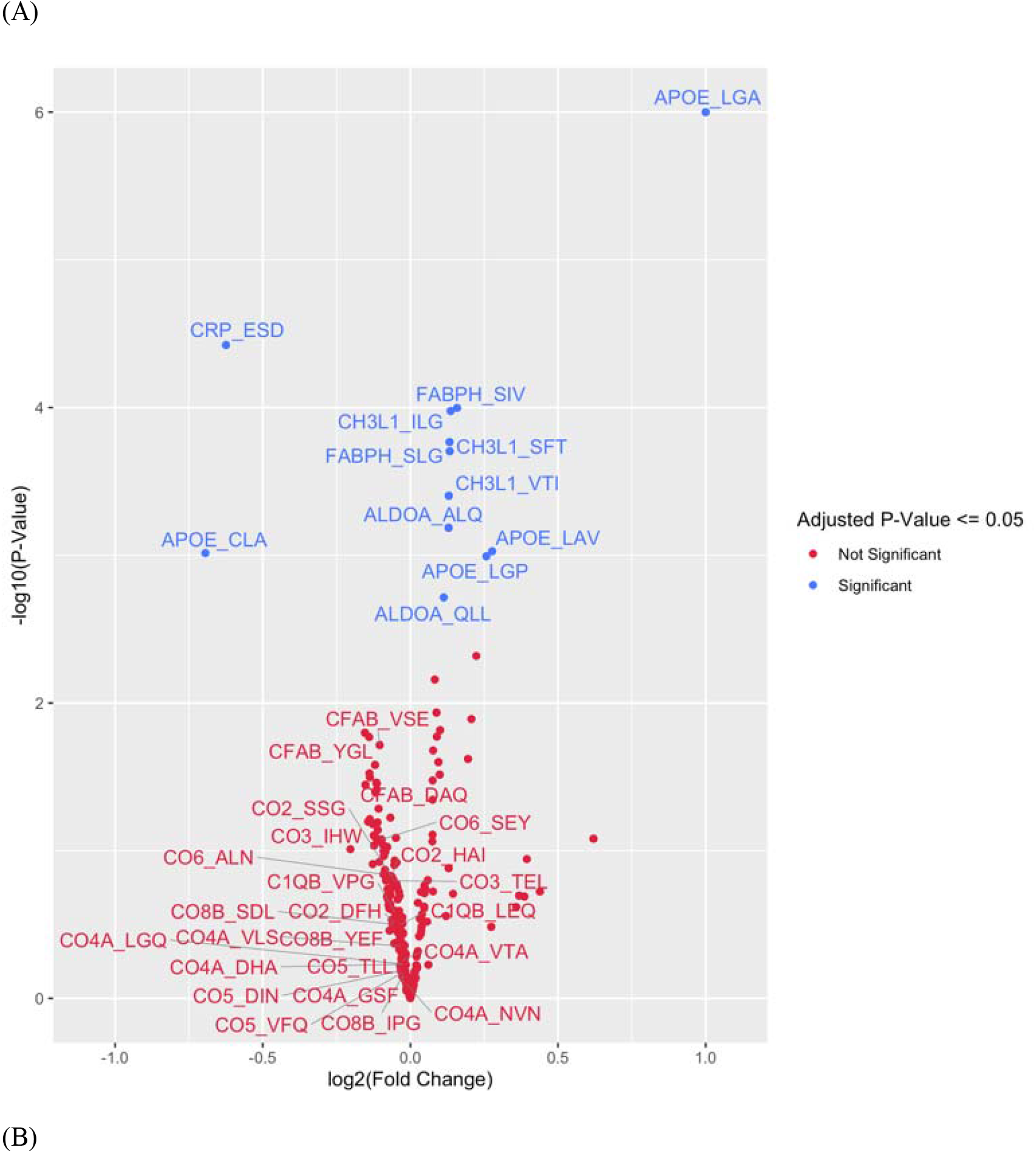
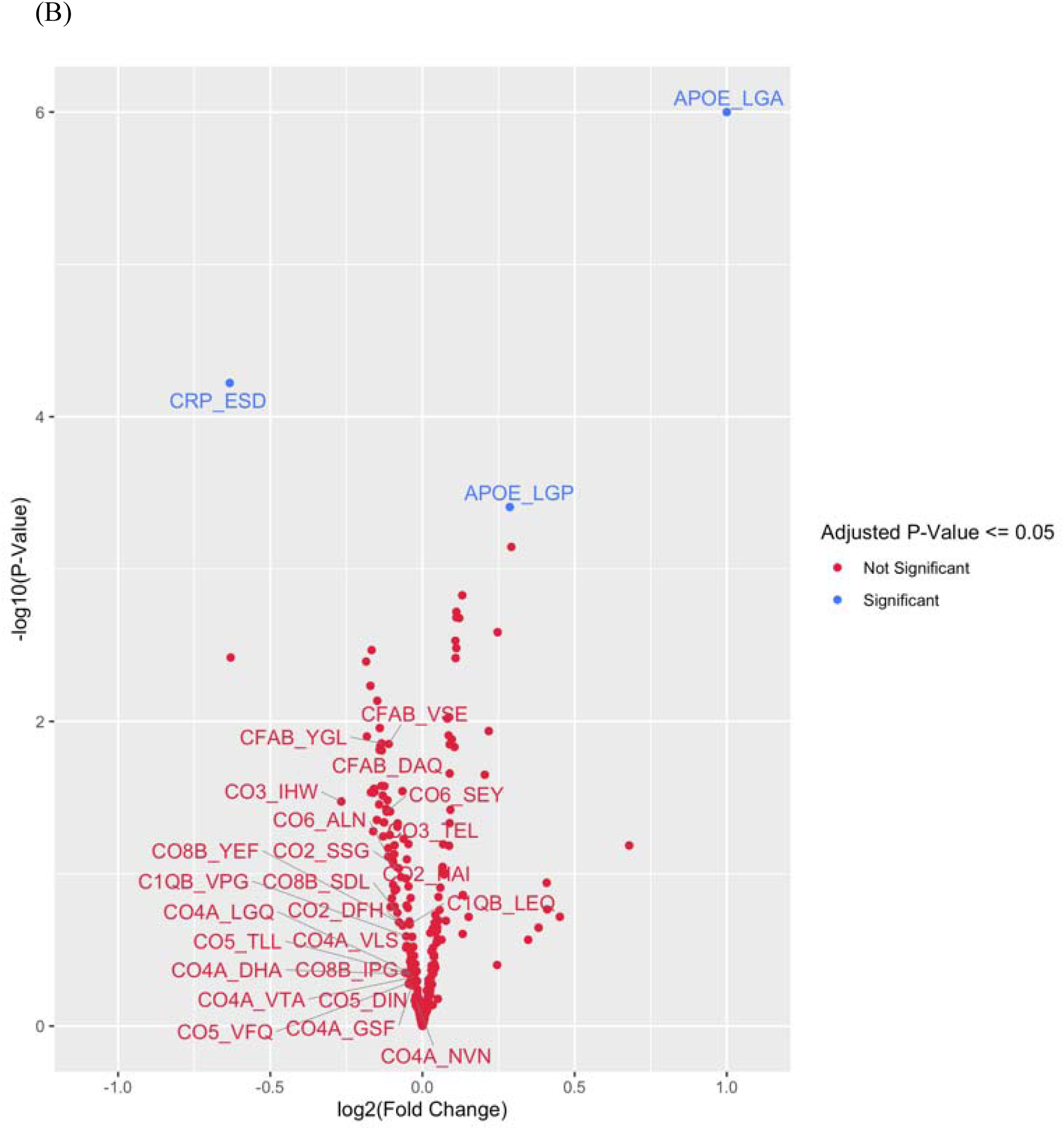
Volcano Plot of CSF Protein/Peptide Expression by APOE genotype, for the top 20 proteins and the complement cascade proteins in model 1 (A), and for the top 20 proteins and the complement cascade proteins in model 2 (B).

All 24 peptides from 8 complement pathway proteins measured in this dataset showed a trend towards lower CSF expression of as a function of increasing *APOE4* gene dose (Table 4, Figure 1A). Although these effects were not significant for any individual complement protein derived peptide (*p* > 0.05 for each, prior to multiple correction comparison), the odds of this happening for 24 unrelated peptides by chance would be 1 over 2^24^, or less than 1 in 16 million. Alternatively, since these 24 peptides were derived from 8 complement pathway proteins, the odds of 8 proteins at random all showing lower expression trends as a function of *APOE4* allele copy number would be 1/2^8^, or a 1 out of 256 chance.

**Table 4:**
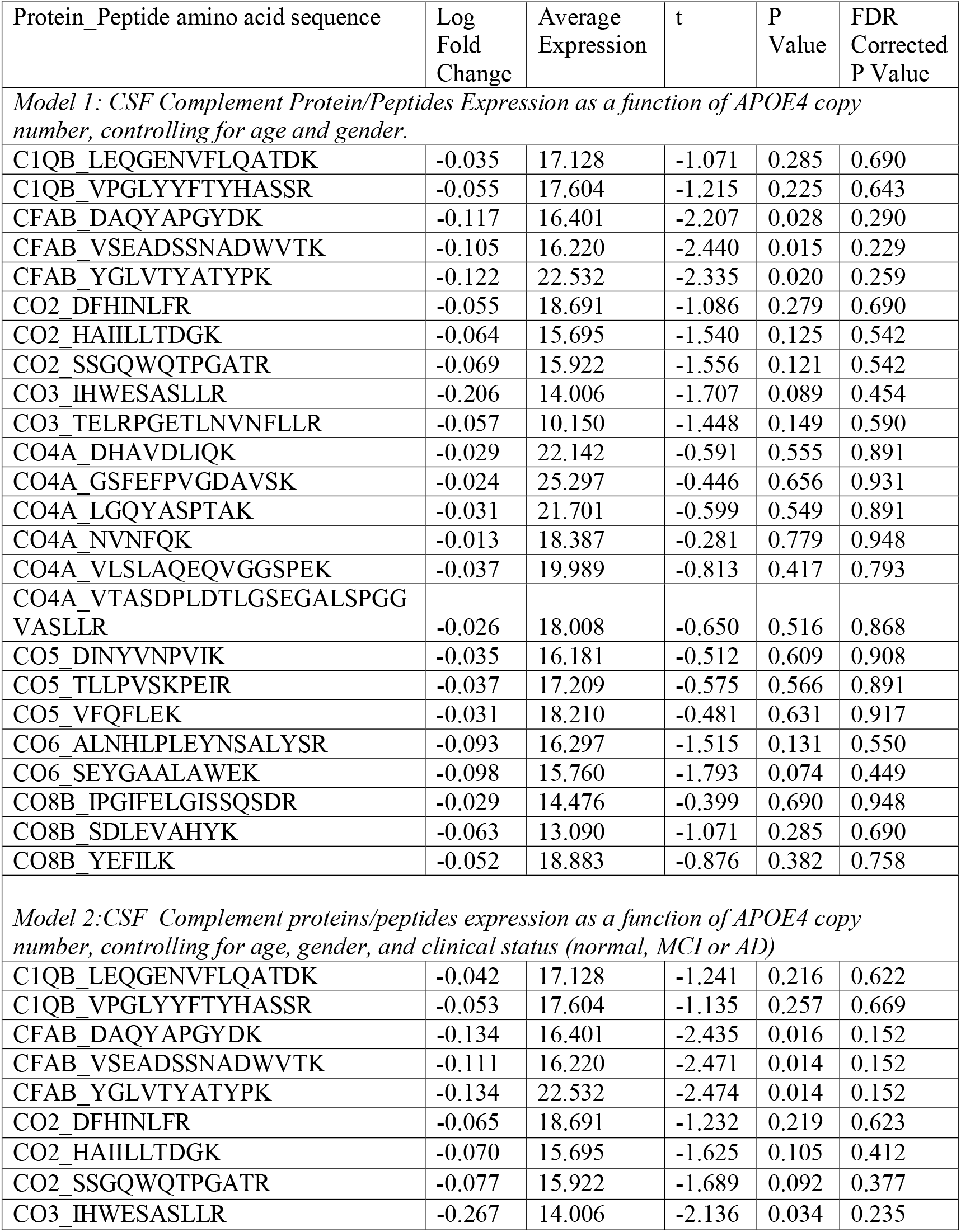
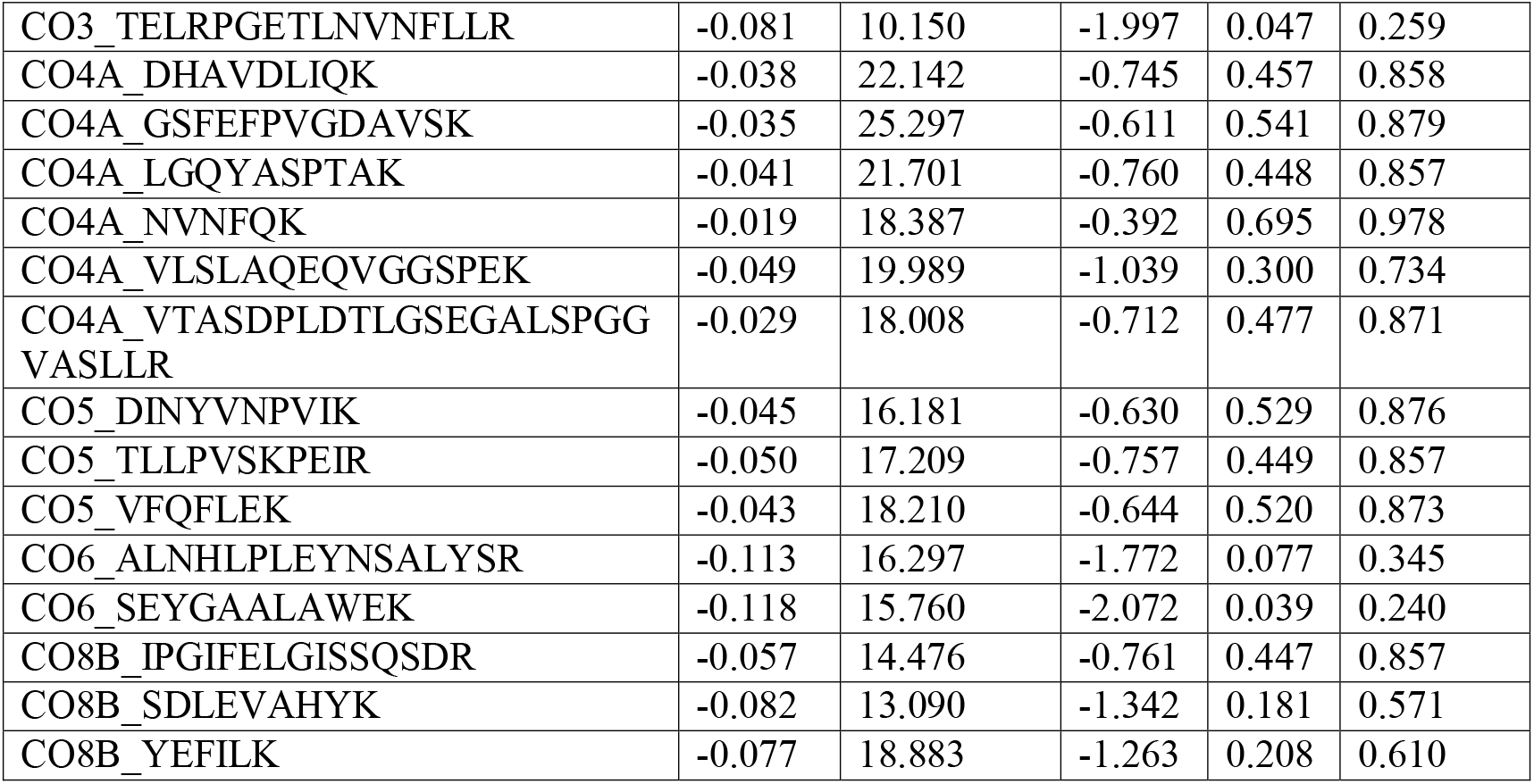
CSF Complement Cascade Peptides/protein Expression by *APOE4* copy number in multivariate models accounting for age and gender (model 1) or age, gender and clinical status (Model 2).

### Model 2: APOE4-dependent CSF peptide changes and clinical status

Because *APOE4* is found in AD patients at disproportionately high frequencies compared to the general population, it is possible that the above findings reflect confounding by AD clinical status (and neurodegeneration) rather than changes directly related to increased *APOE4* copy number itself. Therefore, a second linear model was used that corrected for clinical status (normal control, MCI, or dementia due to AD) in addition to the items in model 1, to test for associations between *APOE4* copy number and CSF peptide expression levels. In this second model, only 3 of 294 peptides had statistically significant *APOE4* copy number-related changes in CSF expression levels (Table 3 and Figure 1B). Increasing APOE4-copy number was associated with increased expression of the *APOE4*-specific peptide (LGADMEDVR) (*q* < 0.01) and decreased expression the CRP-derived peptide (ESDTSYVSLK) (*q*< 0.01). A pan-APOE peptide (LGPLVEQGR) had increased expression associated with increased *APOE4* copy number in this model (*q* = 0.038). CH3L1 (YKL-40)-, FABPH-, and ALDOA-derived peptides that showed significant *APOE4*-copy number-related changes in expression in model 1 (above) no longer remained statistically significant after correcting for disease status and multiple comparisons, although there was still a trend toward increased CSF protein expression for each (*q* = 0.077, q = 0.077, and q = 0.079, respectively). As in the first model (not controlling for AD clinical status), none of the 24 complement protein-derived peptides demonstrated statistically significant differences as a function of *APOE4* copy number. Yet, as in the first model, in this model each of the 24 peptides from the 8 complement pathway-related proteins in this dataset showed trends towards decreasing CSF expression with increasing *APOE4* gene dosage (Table 4). Although none of these trends were statistically significant on their own (p>0.05 for each, prior to multiple comparison) the odds of 24 unrelated peptides all showing this pattern of decreased expression by chance would be less than 1 over 2^24^, or less than 1 in 16 million. Alternatively, since these 24 peptides were derived from 8 complement pathway proteins, the odds of 8 proteins all showing lower expression trends as a function of *APOE4* allele copy number would be 1/2^8^, or a 1 out of 256 chance.

## Discussion

Here, we found that increasing *APOE4* copy number is associated with increased CSF ALDOA, CH3L1 and FABH levels and decreased CSF CRP levels. Further, the CRP decrease remained significant even after controlling for AD clinical status. We also found significant associations between *APOE*4 copy number and several peptides from the APOE protein itself. As expected, we found a strong positive correlation between *APOE4* copy number and CSF expression of the *APOE4* allele specific peptide LGADMEDVR [37, 38]. Both of our statistical models (controlling for age and sex, or for age, sex *and* AD clinical status) showed that increased *APOE4* copy number was associated with increased expression of peptides common to all APOE isoforms, such as LGPLVEQGR, which is often used as a measure of total APOE protein level.

There is conflicting evidence in the literature on whether the *APOE4* carriers have altered CSF APOE protein levels. One study used ELISA assays and found that *APOE4* carriers had higher CSF APOE levels vs non-carriers [39]. Two different studies using mass spectroscopy found no change in CSF APOE levels in *APOE4* carriers [40, 41], though one of them found that *APOE4* carriers had lower plasma APOE levels [40]. Similarly, another study using ELISA assays found reduced peripheral plasma APOE levels in *APOE4* carriers versus non-carriers [42]. Yet, both models in this study showed that increasing *APOE4* copy number was associated with increased CSF pan-APOE peptide levels. It is unclear whether these *APOE4* copy number related increases in APOE protein levels are responsible for increased AD risk, versus whether the increased AD risk is due to functional changes in the APOE protein due to the two amino acid changes encoded by the *APOE4* allele. Indeed, it remains debated in the field to what extent *APOE4*–related increased AD risk is due to a toxic gain of function(s) or a loss of protective function(s) (reviewed in [43]).

We also found that increasing *APOE4* copy number was associated with reduced CSF levels of the CRP-derived peptide (ESDTSYVSLK). CRP is often thought of as a serum biomarker used to follow the acute progression of inflammation and infection [44]. CRP is also increasingly recognized as an active mediator of inflammatory and apoptotic processes, including the activation of the classical complement pathway [45] and the opsonization of atherosclerotic plaques [46] and infarcted myocardial tissues [47]. While elevated CRP levels are typically viewed as an acute marker of active inflammation, low and low-normal CRP levels have been found in chronic inflammatory diseases such as lupus [48], rheumatoid arthritis [49], and inflammatory bowel disease [50, 51]. Thus, the reduced CSF CRP levels observed here may similarly reflect chronically increased inflammation within the CNS of *APOE4* carriers.

Indeed, several studies have consistently found that reduced CRP levels in peripheral blood [52-55] and in CSF [56, 57] correlate with increased cognitive dysfunction and further AD progression in an *APOE4*-dependent manner. Notably, CRP has been implicated in the early development of amyloid plaque formation, neuronal damage, and AD risk [58-61]. The decreased CSF CRP levels observed here may reflect CRP deposition in beta-amyloid plaques and its consumption as a pro-inflammatory mediator in AD pathology, as has been suggested previously [62]. Future studies should examine the role of *APOE4*-dependent modulation of CRP expression and function in AD progression.

We also found that increasing *APOE4* copy was also associated with increased expression of peptides derived from glycoprotein Chitinase 3-like protein 1 (CH3L1; also known as YKL-40) (q< 0.05), the cardiac injury biomarker heart-type fatty acid binding protein (FABPH) (q< 0.05) and the enzyme fructose-bisphosphate aldolase A (ALDOA; q< 0.05) as seen in a prior studies [63]. CH3L1 is a glycoprotein hypothesized to modulate tissue remodeling, and is highly expressed in reactive astrocytes after both acute and chronic neuroinflammation [64-66]. Another recent study using ELISA assays also found increased CSF CH3L1 (YKL-40) levels in *APOE4* carriers [67], further corroborating the findings presented here. Taken together, these findings strongly suggest that *APOE4* carriers have increased neuroinflammation and astroglial activation [68-70] independent of their AD clinical status.

Our finding of *APOE4*-copy number related increases in CSF FABPH levels fits with prior work showing that *APOE4* knock-in mice have elevated hepatic FABPH levels compared to *APOE2* knock-in mice [71]. FABPH is thought to be a potential marker of dyslipidemia that affects membrane stability and contributes to neuronal degeneration as well as atherosclerosis [72], and has been used as a cardiac injury biomarker [73]. While there is strong evidence to link FABPH expression to neuronal loss and AD [74], it is unclear how *APOE4* modulates FABPH expression. It is also unclear whether the *APOE4* copy number related increases in CSF FABPH levels reflect increases in FABPH transcription/translation (or reductions in its breakdown) specifically within the brain, versus within the liver [71] or other peripheral organs.

Another protein that was present at higher levels in the CSF as a function of *APOE4* copy number in this study was ALDOA, a glycolytic enzyme that catalyzes the breakdown of fructose 1-6-diphosphate. Some studies have suggested brain glucose dysregulation plays a key role in Alzheimer’s disease [75], and a recent study has proposed that CSF ALDOA levels are a sensitive and specific biomarker of cognitive impairment due to Alzheimer’s disease [76]. Thus, our finding of an *APOE4* copy number-dependent increase in CSF ALDOA levels may reflect glycolytic dysregulation within the CNS of *APOE4* carriers, which may contribute to AD risk.

The elevations in ALDOA-, FABPH-, and CH3L1 (YKL-40)-derived peptides observed here in model 1 did not remain statistically significant after correcting for AD clinical status, although there remained a trend toward increased CSF protein expression for each even after multiple comparison correction (q = 0.079, q = 0.077, and q = 0.077, respectively). Thus, both *APOE4* copy number and AD clinical status may be associated with increased CSF levels of these three proteins, and the lack of statistical significance in model 2 (accounting for AD clinical status) may be a type statistical 2 error (i.e. insufficient sample size). Future studies with a larger sample size will be necessary to determine the relationship between *APOE4* copy number and CSF levels of these proteins after correcting for AD clinical status.

Further, while not statistically significant, increasing *APOE4* copy number was associated with a trend towards lower expression for all peptides (N=24 total) derived from the 8 complement pathway proteins in this dataset (Fig 1A, B). These trends toward lower complement protein-derived peptide expression in *APOE4* carriers are supported by two other recent studies that also found lower CSF complement protein levels in *APOE4* carriers [77, 78]. Further, these *APOE4* copy number-related trends toward lower CSF complement protein levels were present even after controlling for AD clinical status in model 2. This suggests that *APOE4* copy number may be directly associated with decreased CSF complement protein levels, i.e. that this relationship is not simply due to confounding related to an increased frequency of dementia due to AD in *APOE4* carriers. As discussed above, the chance that the level of 24 peptides (or 8 proteins) would all decrease due to chance alone is extremely low, suggesting that this likely represents a true biological finding. Nonetheless, the average *APOE4* copy number dependent log expression changes for each individual complement protein-derived peptide were small, and would correspond to an absolute reductions of ∼13% and ∼26% in each complement pathway protein in *APOE4* heterozygotes and *APOE4* homozygotes (vs non-carriers), respectively. These lower complement protein levels could represent either decreased transcription/translation or increased degradation in *APOE4* carriers. The former possibility is unlikely, though, because prior work has shown that *APOE4* is not associated with alterations in the transcription or translation of complement pathway proteins [79]. Thus, the findings reported here are most likely consistent with a trend towards increased complement pathway protein degradation in association with increased *APOE4* copy number.

Complement protein degradation can be caused by complement cascade activation, which involves cleavage and degradation of complement proteins [80]. Thus, the trend towards *APOE4-*copy number related reductions in CSF complement protein levels may be a sign of increased complement pathway activation causing increased complement protein degradation in the CNS of *APOE4* carriers. Recent work has suggested that the APOE protein is a negative regulator of complement pathway activation [81]. Taken together with our results, this raises the possibility that the *APOE4* allele results in reduced inhibition (i.e. disinhibition) of the complement pathway, thus resulting in *APOE4* copy number-dependent trends towards lower CSF levels of complement pathway proteins.

Complement-dependent synaptic phagocytosis is thought to represent a neurodegeneration mechanism in AD [82, 83]; thus, our results raise the possibility that *APOE4* may contribute to AD risk by increasing complement pathway activation and resultant synaptic phagocytosis and neurodegeneration. Overall, even though the *APOE4*-related reduction trends in CSF complement protein levels seen here were not statistically significant, the convergence of our human findings with data from cellular [79] and mouse models [84] suggest that further studies are warranted on the relationship between *APOE4* copy number and CSF complement protein levels.

This work has several limitations. First, the data analyzed here were originally obtained to study CSF proteomic correlates of dementia due to AD or MCI [29], rather than to study the effects of *APOE4* copy number on the CSF proteome. Although we controlled for clinical status in model 2, the relatively smaller number of study patients within each clinical status cohort (i.e. normal, MCI, or dementia due to AD) likely limited statistical power to detect effects of *APOE4* on the CSF proteome itself. Thus, future studies on this topic should focus on larger clinically homogenous study populations (i.e. all cognitively normal individuals, or all individuals with dementia due to AD) to reduce variance within each genotype group, and to improve statistical power. Second, the data reported here were from a targeted proteomic platform that only measured 8 of the over 30 proteins in the full complement pathway [85]. Future studies should focus on quantitating the CSF levels of each complement pathway protein to develop a more complete understanding of the relationship between *APOE4* allele copy number and the classical, lectin and alternative complement cascades.

Nonetheless, the data presented here provide strong support for studying the hypothesis that the increased AD risk in *APOE4* carriers is related to early molecular/cellular changes within CRP-related biological processes, and those involving ALDOA-, FABPH- and YKL-40 and the complement pathway.

## Data Availability

Data used in preparation of this article were obtained from the Alzheimer's Disease. Neuroimaging Initiative (ADNI) database (adni.loni.usc.edu). As such, the investigators within the ADNI contributed to the design and implementation of ADNI and/or provided data but did not participate in analysis or writing of this report. A complete listing of ADNI investigators can be found at: http://adni.loni.usc.edu/wp-content/uploads/how_to_apply/ADNI_ Acknowledgement_List.pdf
Data collection and sharing for this project was funded by the Alzheimer's Disease Neuroimaging Initiative (ADNI) (National Institutes of Health Grant U01 AG024904) and DOD ADNI (Department of Defense award number W81XWH-12-2-0012). ADNI is funded by the National Institute on Aging, the National Institute of Biomedical Imaging and Bioengineering, and through generous contributions from the following: AbbVie, Alzheimer's Association; Alzheimer's Drug Discovery Foundation; Araclon Biotech; BioClinica, Inc.; Biogen; Bristol-Myers Squibb Company; CereSpir, Inc.; Cogstate; Eisai Inc.; Elan Pharmaceuticals, Inc.; Eli Lilly and Company; EuroImmun; F. Hoffmann-La Roche Ltd and its affiliated company Genentech, Inc.; Fujirebio; GE Healthcare; IXICO Ltd.; Janssen Alzheimer Immunotherapy Research & Development, LLC.; Johnson & Johnson Pharmaceutical Research & Development LLC.; Lumosity; Lundbeck; Merck & Co., Inc.; Meso Scale Diagnostics, LLC.; NeuroRx Research; Neurotrack Technologies; Novartis Pharmaceuticals Corporation; Pfizer Inc.; Piramal Imaging; Servier; Takeda Pharmaceutical Company; and Transition Therapeutics. The Canadian Institutes of Health Research is providing funds to support ADNI clinical sites in Canada. Private sector contributions are facilitated by the Foundation for the National Institutes of Health (www.fnih.org). The grantee organization is the Northern California Institute for Research and Education, and the study is coordinated by the Alzheimer's Therapeutic Research Institute at the University of Southern California. ADNI data are disseminated by the Laboratory for Neuro Imaging at the University of Southern California.

## Acknowledgements

Data used in preparation of this article were obtained from the Alzheimer’s Disease. Neuroimaging Initiative (ADNI) database (adni.loni.usc.edu). As such, the investigators within the ADNI contributed to the design and implementation of ADNI and/or provided data but did not participate in analysis or writing of this report. A complete listing of ADNI investigators can be found at: http://adni.loni.usc.edu/wp-content/uploads/how_to_apply/ADNI_Acknowledgement_List.pdf

Data collection and sharing for this project was funded by the Alzheimer’s Disease Neuroimaging Initiative (ADNI) (National Institutes of Health Grant U01 AG024904) and DOD ADNI (Department of Defense award number W81XWH-12-2-0012). ADNI is funded by the National Institute on Aging, the National Institute of Biomedical Imaging and Bioengineering, and through generous contributions from the following: AbbVie, Alzheimer’s Association; Alzheimer’s Drug Discovery Foundation; Araclon Biotech; BioClinica, Inc.; Biogen; Bristol-Myers Squibb Company; CereSpir, Inc.; Cogstate; Eisai Inc.; Elan Pharmaceuticals, Inc.; Eli Lilly and Company; EuroImmun; F. Hoffmann-La Roche Ltd and its affiliated company

Genentech, Inc.; Fujirebio; GE Healthcare; IXICO Ltd.; Janssen Alzheimer Immunotherapy Research & Development, LLC.; Johnson & Johnson Pharmaceutical Research & Development LLC.; Lumosity; Lundbeck; Merck & Co., Inc.; Meso Scale Diagnostics, LLC.; NeuroRx Research; Neurotrack Technologies; Novartis Pharmaceuticals Corporation; Pfizer Inc.; Piramal Imaging; Servier; Takeda Pharmaceutical Company; and Transition Therapeutics. The Canadian Institutes of Health Research is providing funds to support ADNI clinical sites in Canada. Private sector contributions are facilitated by the Foundation for the National Institutes of Health (www.fnih.org). The grantee organization is the Northern California Institute for Research and Education, and the study is coordinated by the Alzheimer’s Therapeutic Research Institute at the University of Southern California. ADNI data are disseminated by the Laboratory for Neuro Imaging at the University of Southern California.

## Notes

### Competing Interest Statement

Dr. Berger acknowledges consulting income from two legal cases regarding postoperative cognitive change.

### Funding Statement

Dr. Berger acknowledges support from K76-AG057022, and additional support from NIH grants P30AG028716 and UH2AG056925; a program to advance clinical trials (PACT) grant from the Alzheimer's Drug Discovery Foundation, the inaugural Ann Bussel award from the Ruth K. Broad Foundation at Duke University, and the Duke Anesthesiology Department. Dr. Devinney acknowledges support from a research fellowship grant from the Foundation for Anesthesia Education and Research.

## References

[1] Corder E, Saunders A, Strittmatter W, Schmechel D, Gaskell P, Small G, Roses A, Haines J, Pericak-Vance M (1993) Gene dose of apolipoprotein E type 4 allele and the risk of Alzheimer’s disease in late onset families. Science 261, 921–923.

[2] Neu SC, Pa J, Kukull W, Beekly D, Kuzma A, Gangadharan P, Wang LS, Romero K, Arneric SP, Redolfi A, Orlandi D, Frisoni GB, Au R, Devine S, Auerbach S, Espinosa A, Boada M, Ruiz A, Johnson SC, Koscik R, Wang JJ, Hsu WC, Chen YL, Toga AW (2017) Apolipoprotein E Genotype and Sex Risk Factors for Alzheimer Disease: A Meta-analysis. JAMA Neurol 74, 1178–1189.

[3] Loy CT, Schofield PR, Turner AM, Kwok JBJ (2014) Genetics of dementia. The Lancet 383, 828–840.

[4] Holtzman DM, Herz J, Bu G (2012) Apolipoprotein E and apolipoprotein E receptors: normal biology and roles in Alzheimer disease. Cold Spring Harb Perspect Med 2, a006312.

[5] Michaelson DM (2014) APOE ε4: The most prevalent yet understudied risk factor for Alzheimer’s disease. Alzheimer’s & Dementia 10, 861–868.

[6] Fazekas F, Strasser–Fuchs S, Kollegger H, Berger T, Kristoferitsch W, Schmidt H, Enzinger C, Schiefermeier M, Schwarz C, Kornek B, Reindl M, Huber K, Grass R, Wimmer G, Vass K, Pfeiffer KH, Hartung HP, Schmidt R (2001) Apolipoprotein E ε4 is associated with rapid progression of multiple sclerosis. Neurology 57, 853–857.

[7] Yue JK, Robinson CK, Burke JF, Winkler EA, Deng H, Cnossen MC, Lingsma HF, Ferguson AR, McAllister TW, Rosand J, Burchard EG, Sorani MD, Sharma S, Nielson JL, Satris GG, Talbott JF, Tarapore PE, Korley FK, Wang KKW, Yuh EL, Mukherjee P, Diaz-Arrastia R, Valadka AB, Okonkwo DO, Manley GT, Investigators T-T (2017) Apolipoprotein E epsilon 4 (APOE-epsilon4) genotype is associated with decreased 6-month verbal memory performance after mild traumatic brain injury. Brain Behav 7, e00791.

[8] Martinez-Gonzalez NA, Sudlow CL (2006) Effects of apolipoprotein E genotype on outcome after ischaemic stroke, intracerebral haemorrhage and subarachnoid haemorrhage. J Neurol Neurosurg Psychiatry 77, 1329–1335.

[9] Mahley RW (2016) Apolipoprotein E: from cardiovascular disease to neurodegenerative disorders. J Mol Med (Berl) 94, 739–746.

[10] Kulminski AM, Arbeev KG, Culminskaya I, Arbeeva L, Ukraintseva SV, Stallard E, Christensen K, Schupf N, Province MA, Yashin AI (2014) Age, gender, and cancer but not neurodegenerative and cardiovascular diseases strongly modulate systemic effect of the Apolipoprotein E4 allele on lifespan. PLoS Genet 10, e1004141.

[11] Deelen J, Evans DS, Arking DE, Tesi N, Nygaard M, Liu X, Wojczynski MK, Biggs ML, van der Spek A, Atzmon G, Ware EB, Sarnowski C, Smith AV, Seppala I, Cordell HJ, Dose J, Amin N, Arnold AM, Ayers KL, Barzilai N, Becker EJ, Beekman M, Blanche H, Christensen K, Christiansen L, Collerton JC, Cubaynes S, Cummings SR, Davies K, Debrabant B, Deleuze JF, Duncan R, Faul JD, Franceschi C, Galan P, Gudnason V, Harris TB, Huisman M, Hurme MA, Jagger C, Jansen I, Jylha M, Kahonen M, Karasik D, Kardia SLR, Kingston A, Kirkwood TBL, Launer LJ, Lehtimaki T, Lieb W, Lyytikainen LP, Martin-Ruiz C, Min J, Nebel A, Newman AB, Nie C, Nohr EA, Orwoll ES, Perls TT, Province MA, Psaty BM, Raitakari OT, Reinders MJT, Robine JM, Rotter JI, Sebastiani P, Smith J, Sorensen TIA, Taylor KD, Uitterlinden AG, van der Flier W, van der Lee SJ, van Duijn CM, van Heemst D, Vaupel JW, Weir D, Ye K, Zeng Y, Zheng W, Holstege H, Kiel DP, Lunetta KL, Slagboom PE, Murabito JM (2019) A meta-analysis of genome-wide association studies identifies multiple longevity genes. Nat Commun 10, 3669.

[12] Yamazaki Y, Zhao N, Caulfield TR, Liu CC, Bu G (2019) Apolipoprotein E and Alzheimer disease: pathobiology and targeting strategies. Nat Rev Neurol 15, 501–518.

[13] Iacono D, Feltis G Impact of Apolipoprotein E gene polymorphism during normal and pathological conditions of the brain across the lifespan. Aging 11, 787–816.

[14] Ghisays V, Goradia DD, Protas H, Bauer RJ, 3rd, Devadas V, Tariot PN, Lowe VJ, Knopman DS, Petersen RC, Jack CR, Jr., Caselli RJ, Su Y, Chen K, Reiman EM (2020) Brain imaging measurements of fibrillar amyloid-beta burden, paired helical filament tau burden, and atrophy in cognitively unimpaired persons with two, one, and no copies of the APOE epsilon4 allele. Alzheimers Dement 16, 598–609.

[15] Jansen WJ, Ossenkoppele R, Knol DL, Tijms BM, Scheltens P, Verhey FR, Visser PJ, Amyloid Biomarker Study G, Aalten P, Aarsland D, Alcolea D, Alexander M, Almdahl IS, Arnold SE, Baldeiras I, Barthel H, van Berckel BN, Bibeau K, Blennow K, Brooks DJ, van Buchem MA, Camus V, Cavedo E, Chen K, Chetelat G, Cohen AD, Drzezga A, Engelborghs S, Fagan AM, Fladby T, Fleisher AS, van der Flier WM, Ford L, Forster S, Fortea J, Foskett N, Frederiksen KS, Freund-Levi Y, Frisoni GB, Froelich L, Gabryelewicz T, Gill KD, Gkatzima O, Gomez-Tortosa E, Gordon MF, Grimmer T, Hampel H, Hausner L, Hellwig S, Herukka SK, Hildebrandt H, Ishihara L, Ivanoiu A, Jagust WJ, Johannsen P, Kandimalla R, Kapaki E, Klimkowicz-Mrowiec A, Klunk WE, Kohler S, Koglin N, Kornhuber J, Kramberger MG, Van Laere K, Landau SM, Lee DY, de Leon M, Lisetti V, Lleo A, Madsen K, Maier W, Marcusson J, Mattsson N, de Mendonca A, Meulenbroek O, Meyer PT, Mintun MA, Mok V, Molinuevo JL, Mollergard HM, Morris JC, Mroczko B, Van der Mussele S, Na DL, Newberg A, Nordberg A, Nordlund A, Novak GP, Paraskevas GP, Parnetti L, Perera G, Peters O, Popp J, Prabhakar S, Rabinovici GD, Ramakers IH, Rami L, Resende de Oliveira C, Rinne JO, Rodrigue KM, Rodriguez-Rodriguez E, Roe CM, Rot U, Rowe CC, Ruther E, Sabri O, Sanchez-Juan P, Santana I, Sarazin M, Schroder J, Schutte C, Seo SW, Soetewey F, Soininen H, Spiru L, Struyfs H, Teunissen CE, Tsolaki M, Vandenberghe R, Verbeek MM, Villemagne VL, Vos SJ, van Waalwijk van Doorn LJ, Waldemar G, Wallin A, Wallin AK, Wiltfang J, Wolk DA, Zboch M, Zetterberg H (2015) Prevalence of cerebral amyloid pathology in persons without dementia: a meta-analysis. JAMA 313, 1924–1938.

[16] Hodgetts CJ, Shine JP, Williams H, Postans M, Sims R, Williams J, Lawrence AD, Graham KS (2019) Increased posterior default mode network activity and structural connectivity in young adult APOE-epsilon4 carriers: a multimodal imaging investigation. Neurobiol Aging 73, 82–91.

[17] Dennis NA, Browndyke JN, Stokes J, Need A, Burke JR, Welsh-Bohmer KA, Cabeza R (2010) Temporal lobe functional activity and connectivity in young adult APOE lll4 carriers. Alzheimer’s & Dementia 6, 303–311.

[18] Molinuevo JL, Ayton S, Batrla R, Bednar MM, Bittner T, Cummings J, Fagan AM, Hampel H, Mielke MM, Mikulskis A, O’Bryant S, Scheltens P, Sevigny J, Shaw LM, Soares HD, Tong G, Trojanowski JQ, Zetterberg H, Blennow K (2018) Current state of Alzheimer’s fluid biomarkers. Acta Neuropathol 136, 821–853.

[19] Zhu Y, Nwabuisi-Heath E, Dumanis SB, Tai LM, Yu C, Rebeck GW, LaDu MJ (2012) APOE genotype alters glial activation and loss of synaptic markers in mice. Glia 60, 559–569.

[20] Vitek MP, Brown CM, Colton CA (2009) APOE genotype-specific differences in the innate immune response. Neurobiol Aging 30, 1350–1360.

[21] Tsoi L-M, Wong K-Y, Liu Y-M, Ho Y-Y (2007) Apoprotein E isoform-dependent expression and secretion of pro-inflammatory cytokines TNF-α and IL-6 in macrophages. Archives of Biochemistry and Biophysics 460, 33–40.

[22] Weiner MW, Aisen PS, Jack CR, Jr., Jagust WJ, Trojanowski JQ, Shaw L, Saykin AJ, Morris JC, Cairns N, Beckett LA, Toga A, Green R, Walter S, Soares H, Snyder P, Siemers E, Potter W, Cole PE, Schmidt M, Alzheimer’s Disease Neuroimaging I (2010) The Alzheimer’s disease neuroimaging initiative: progress report and future plans. Alzheimers Dement 6, 202–211 e207.

[23] Arevalo-Rodriguez I, Smailagic N, Roque IFM, Ciapponi A, Sanchez-Perez E, Giannakou A, Pedraza OL, Bonfill Cosp X, Cullum S (2015) Mini-Mental State Examination (MMSE) for the detection of Alzheimer’s disease and other dementias in people with mild cognitive impairment (MCI). Cochrane Database Syst Rev, CD010783.

[24] Morris JC (1997) Clinical Dementia Rating: A Reliable and Valid Diagnostic and Staging Measure for Dementia of the Alzheimer Type. Int. Psychogeriatr. 9, 173.

[25] Wechsler D (1987) WMS-R: Wechsler Memory Scale-Revised: Manual, Psychological Corporation.

[26] McKhann GM, Knopman DS, Chertkow H, Hyman BT, Jack CR, Jr., Kawas CH, Klunk WE, Koroshetz WJ, Manly JJ, Mayeux R, Mohs RC, Morris JC, Rossor MN, Scheltens P, Carrillo MC, Thies B, Weintraub S, Phelps CH (2011) The diagnosis of dementia due to Alzheimer’s disease: recommendations from the National Institute on Aging-Alzheimer’s Association workgroups on diagnostic guidelines for Alzheimer’s disease. Alzheimers Dement 7, 263–269.

[27] Shaw LM, Vanderstichele H, Knapik-Czajka M, Figurski M, Coart E, Blennow K, Soares H, Simon AJ, Lewczuk P, Dean RA, Siemers E, Potter W, Lee VM, Trojanowski JQ, Alzheimer’s Disease Neuroimaging I (2011) Qualification of the analytical and clinical performance of CSF biomarker analyses in ADNI. Acta Neuropathol 121, 597–609.

[28] Shaw LM, Vanderstichele H, Knapik-Czajka M, Clark CM, Aisen PS, Petersen RC, Blennow K, Soares H, Simon A, Lewczuk P, Dean R, Siemers E, Potter W, Lee VM, Trojanowski JQ, Alzheimer’s Disease Neuroimaging I (2009) Cerebrospinal fluid biomarker signature in Alzheimer’s disease neuroimaging initiative subjects. Ann Neurol 65, 403–413.

[29] Spellman DS, Wildsmith KR, Honigberg LA, Tuefferd M, Baker D, Raghavan N, Nairn AC, Croteau P, Schirm M, Allard R, Lamontagne J, Chelsky D, Hoffmann S, Potter WZ, Alzheimer’s Disease Neuroimaging I, Foundation for NIHBCCSFPPT (2015) Development and evaluation of a multiplexed mass spectrometry based assay for measuring candidate peptide biomarkers in Alzheimer’s Disease Neuroimaging Initiative (ADNI) CSF. Proteomics Clin Appl 9, 715–731.

[30] Choi YS, Lee KH (2016) Multiple reaction monitoring assay based on conventional liquid chromatography and electrospray ionization for simultaneous monitoring of multiple cerebrospinal fluid biomarker candidates for Alzheimer’s disease. Arch Pharm Res 39, 390–397.

[31] Shaw LM, Arias J, Blennow K, Galasko D, Molinuevo JL, Salloway S, Schindler S, Carrillo MC, Hendrix JA, Ross A, Illes J, Ramus C, Fifer S (2018) Appropriate use criteria for lumbar puncture and cerebrospinal fluid testing in the diagnosis of Alzheimer’s disease. Alzheimers Dement 14, 1505–1521.

[32] Blennow K, Shaw LM, Stomrud E, Mattsson N, Toledo JB, Buck K, Wahl S, Eichenlaub U, Lifke V, Simon M, Trojanowski JQ, Hansson O (2019) Predicting clinical decline and conversion to Alzheimer’s disease or dementia using novel Elecsys Aβ(1-42), pTau and tTau CSF immunoassays. Sci Rep 9, 19024.

[33] Ritchie ME, Phipson B, Wu D, Hu Y, Law CW, Shi W, Smyth GK (2015) limma powers differential expression analyses for RNA-sequencing and microarray studies. Nucleic Acids Res 43, e47.

[34] Gentleman R, Carey V, Bates D, Bolstad B, Dettling M, Dudoit S, Ellis B, Gautier L, Ge Y, Gentry J, Hornik K, Hothorn T, Huber W, Iacus S, Irizarry R, Leisch F, Li C, Maechler M, Rossini A, Sawitzki G, Smith C, Smyth G, Tierney L, Yang J, Zhang J (2004) Bioconductor: open software development for computational biology and bioinformatics. Genome Biol 5, R80.

[35] Millard SP, Lutz F, Li G, Galasko DR, Farlow MR, Quinn JF, Kaye JA, Leverenz JB, Tsuang D, Yu CE, Peskind ER, Bekris LM (2014) Association of cerebrospinal fluid Aβ42 with A2M gene in cognitively normal subjects. Neurobiol Aging 35, 357–364.

[36] Kok E, Haikonen S, Luoto T, Huhtala H, Goebeler S, Haapasalo H, Karhunen PJ (2009) Apolipoprotein E-dependent accumulation of Alzheimer disease-related lesions begins in middle age. Ann Neurol 65, 650–657.

[37] Llano DA, Bundela S, Mudar RA, Devanarayan V, Alzheimer’s Disease Neuroimaging I (2017) A multivariate predictive modeling approach reveals a novel CSF peptide signature for both Alzheimer’s Disease state classification and for predicting future disease progression. PLoS One 12, e0182098.

[38] Simon R, Girod M, Fonbonne C, Salvador A, Clement Y, Lanteri P, Amouyel P, Lambert JC, Lemoine J (2012) Total ApoE and ApoE4 isoform assays in an Alzheimer’s disease case-control study by targeted mass spectrometry (n=669): a pilot assay for methionine-containing proteotypic peptides. Mol Cell Proteomics 11, 1389–1403.

[39] van Harten AC, Jongbloed W, Teunissen CE, Scheltens P, Veerhuis R, van der Flier WM (2017) CSF ApoE predicts clinical progression in nondemented APOEε4 carriers. Neurobiol Aging 57, 186–194.

[40] Mart ínez-Morillo E, Hansson O, Atagi Y, Bu G, Minthon L, Diamandis EP, Nielsen HM (2014) Total apolipoprotein E levels and specific isoform composition in cerebrospinal fluid and plasma from Alzheimer’s disease patients and controls. Acta Neuropathologica 127, 633–643.

[41] Minta K, Brinkmalm G, Janelidze S, Sjödin S, Portelius E, Stomrud E, Zetterberg H, Blennow K, Hansson O, Andreasson U (2020) Quantification of total apolipoprotein E and its isoforms in cerebrospinal fluid from patients with neurodegenerative diseases. Alzheimers Res Ther 12, 19.

[42] Gupta VB, Laws SM, Villemagne VL, Ames D, Bush AI, Ellis KA, Lui JK, Masters C, Rowe CC, Szoeke C, Taddei K, Martins RN (2011) Plasma apolipoprotein E and Alzheimer disease risk. The AIBL study of aging 76, 1091–1098.

[43] Safieh M, Korczyn AD, Michaelson DM (2019) ApoE4: an emerging therapeutic target for Alzheimer’s disease. BMC Medicine 17, 64.

[44] Sproston NR, Ashworth JJ (2018) Role of C-Reactive Protein at Sites of Inflammation and Infection. Front Immunol 9, 754.

[45] Mihlan M, Blom AM, Kupreishvili K, Lauer N, Stelzner K, Bergstrom F, Niessen HW, Zipfel PF (2011) Monomeric C-reactive protein modulates classic complement activation on necrotic cells. FASEB J 25, 4198–4210.

[46] Torzewski J, Torzewski M, Bowyer D, Frohlich M, Koenig W, Waltenberger J, Fitzsimmons C, Hombach V (1998) C-Reactive Protein Frequently Colocalizes With the Terminal Complement Complex in the Intima of Early Atherosclerotic Lesions of Human Coronary Arteries. Arterioscler Thromb Vasc Biol. 18, 1386–1392.

[47] Lagrand W, Niessen H, Wolbink G, Jaspars L, Visser C, Verheugt F, Meijer C, Hack C (1997) C-Reactive Protein Colocalizes With Complement in Human Hearts During Acute Myocardial Infarction. Circulation 95, 97–103.

[48] Suh CH, Chun HY, Ye YM, Park HS (2006) Unresponsiveness of C-reactive protein in the non-infectious inflammation of systemic lupus erythematosus is associated with interleukin 6. Clin Immunol 119, 291–296.

[49] Kay J, Morgacheva O, Messing SP, Kremer JM, Greenberg JD, Reed GW, Gravallese EM, Furst DE (2014) Clinical disease activity and acute phase reactant levels are discordant among patients with active rheumatoid arthritis: acute phase reactant levels contribute separately to predicting outcome at one year. Arthritis Res Ther 16, R40.

[50] Yang DH, Yang SK, Park SH, Lee HS, Boo SJ, Park JH, Na SY, Jung KW, Kim KJ, Ye BD, Byeon JS, Myung SJ (2015) Usefulness of C-reactive protein as a disease activity marker in Crohn’s disease according to the location of disease. Gut Liver 9, 80–86.

[51] Florin TH, Paterson EW, Fowler EV, Radford-Smith GL (2006) Clinically active Crohn’s disease in the presence of a low C-reactive protein. Scand J Gastroenterol 41, 306–311.

[52] Royall DR, Al-Rubaye S, Bishnoi R, Palmer RF (2017) Few serum proteins mediate APOE’s association with dementia. PLoS One 12, e0172268.

[53] Haan MN, Aiello AE, West NA, Jagust WJ (2008) C-reactive protein and rate of dementia in carriers and non carriers of Apolipoprotein APOE4 genotype. Neurobiol Aging 29, 1774–1782.

[54] Hubacek JA, Peasey A, Pikhart H, Stavek P, Kubinova R, Marmot M, Bobak M (2010) APOE polymorphism and its effect on plasma C-reactive protein levels in a large general population sample. Hum Immunol 71, 304–308.

[55] Locascio JJ, Fukumoto H, Yap L, Bottiglieri T, Growdon JH, Hyman BT, Irizarry MC (2008) Plasma amyloid beta-protein and C-reactive protein in relation to the rate of progression of Alzheimer disease. Arch Neurol 65, 776–785.

[56] Brosseron F, Traschütz A, Widmann CN, Kummer MP, Tacik P, Santarelli F, Jessen F, Heneka MT (2018) Characterization and clinical use of inflammatory cerebrospinal fluid protein markers in Alzheimer’s disease. Alzheimers Res Ther 10, 25.

[57] Schuitemaker A, Dik MG, Veerhuis R, Scheltens P, Schoonenboom NS, Hack CE, Blankenstein MA, Jonker C (2009) Inflammatory markers in AD and MCI patients with different biomarker profiles. Neurobiol Aging 30, 1885–1889.

[58] Iwamoto N, Nishiyama E, Ohwada J, Arai H (1994) Demonstration of CRP immunoreactivity in brains of Alzheimer’s disease: immunohistochemical study using formic acid pretreatment of tissue sections. Neurosci Lett 177, 23–26.

[59] Strang F, Scheichl A, Chen YC, Wang X, Htun NM, Bassler N, Eisenhardt SU, Habersberger J, Peter K (2012) Amyloid plaques dissociate pentameric to monomeric C-reactive protein: a novel pathomechanism driving cortical inflammation in Alzheimer’s disease? Brain Pathol 22, 337–346.

[60] Bi BT, Lin HB, Cheng YF, Zhou H, Lin T, Zhang MZ, Li TJ, Xu JP (2012) Promotion of β-amyloid production by C-reactive protein and its implications in the early pathogenesis of Alzheimer’s disease. Neurochem Int 60, 257–266.

[61] Slevin M, Matou S, Zeinolabediny Y, Corpas R, Weston R, Liu D, Boras E, Di Napoli M, Petcu E, Sarroca S, Popa-Wagner A, Love S, Font MA, Potempa LA, Al-Baradie R, Sanfeliu C, Revilla S, Badimon L, Krupinski J (2015) Monomeric C-reactive protein--a key molecule driving development of Alzheimer’s disease associated with brain ischaemia? Sci Rep 5, 13281.

[62] McFadyen JD, Zeller J, Potempa LA, Pietersz GA, Eisenhardt SU, Peter K (2020) C-Reactive Protein and Its Structural Isoforms: An Evolutionary Conserved Marker and Central Player in Inflammatory Diseases and Beyond In Vertebrate and Invertebrate Respiratory Proteins, Lipoproteins and other Body Fluid Proteins Springer International Publishing, pp. 499–520.

[63] Reus LM, Stringer S, Posthuma D, Teunissen CE, Scheltens P, Pijnenburg YAL, Visser PJ, Tijms BM (2020) Degree of genetic liability for Alzheimer’s disease associated with specific proteomic profiles in cerebrospinal fluid. Neurobiol Aging 93, 144.e141-144.e115.

[64] Johansen JS, Baslund B, Garbarsch C, Hansen M, Stoltenberg M, Lorenzen I, Price PA (1999) YKL-40 in giant cells and macrophages from patients with giant cell arteritis. Arthritis Rheum 42, 2624–2630.

[65] Junker N, Johansen JS, Andersen CB, Kristjansen PE (2005) Expression of YKL-40 by peritumoral macrophages in human small cell lung cancer. Lung Cancer 48, 223–231.

[66] Létuvé S, Kozhich A, Arouche N, Grandsaigne M, Reed J, Dombret MC, Kiener PA, Aubier M, Coyle AJ, Pretolani M (2008) YKL-40 is elevated in patients with chronic obstructive pulmonary disease and activates alveolar macrophages. J Immunol 181, 5167–5173.

[67] Wang L, Gao T, Cai T, Li K, Zheng P, Liu J (2020) Cerebrospinal fluid levels of YKL-40 in prodromal Alzheimer’s disease. Neurosci Lett 715, 134658.

[68] Bonneh-Barkay D, Bissel SJ, Kofler J, Starkey A, Wang G, Wiley CA (2012) Astrocyte and macrophage regulation of YKL-40 expression and cellular response in neuroinflammation. Brain Pathol 22, 530–546.

[69] Bonneh-Barkay D, Wang G, Starkey A, Hamilton RL, Wiley CA (2010) In vivo CHI3L1 (YKL-40) expression in astrocytes in acute and chronic neurological diseases. J Neuroinflammation 7, 34.

[70] Wiley CA, Bonneh-Barkay D, Dixon CE, Lesniak A, Wang G, Bissel SJ, Kochanek PM (2015) Role for mammalian chitinase 3-like protein 1 in traumatic brain injury. Neuropathology 35, 95–106.

[71] Conway V, Larouche A, Alata W, Vandal M, Calon F, Plourde M (2014) Apolipoprotein E isoforms disrupt long-chain fatty acid distribution in the plasma, the liver and the adipose tissue of mice. Prostaglandins Leukot Essent Fatty Acids 91, 261–267.

[72] Bjerke M, Kern S, Blennow K, Zetterberg H, Waern M, Borjesson-Hanson A, Ostling S, Kern J, Skoog I (2016) Cerebrospinal Fluid Fatty Acid-Binding Protein 3 is Related to Dementia Development in a Population-Based Sample of Older Adult Women Followed for 8 Years. J Alzheimers Dis 49, 733–741.

[73] Otaki Y, Watanabe T, Kubota I (2017) Heart-type fatty acid-binding protein in cardiovascular disease: A systemic review. Clin Chim Acta 474, 44–53.

[74] Desikan RS, Thompson WK, Holland D, Hess CP, Brewer JB, Zetterberg H, Blennow K, Andreassen OA, McEvoy LK, Hyman BT, Dale AM (2013) Heart fatty acid binding protein and Aβ-associated Alzheimer’s neurodegeneration. Mol Neurodegener 8, 39.

[75] An Y, Varma VR, Varma S, Casanova R, Dammer E, Pletnikova O, Chia CW, Egan JM, Ferrucci L, Troncoso J, Levey AI, Lah J, Seyfried NT, Legido-Quigley C, O’Brien R, Thambisetty M (2018) Evidence for brain glucose dysregulation in Alzheimer’s disease. Alzheimers Dement 14, 318–329.

[76] Zhou M, Haque RU, Dammer EB, Duong DM, Ping L, Johnson ECB, Lah JJ, Levey AI, Seyfried NT (2020) Targeted mass spectrometry to quantify brain-derived cerebrospinal fluid biomarkers in Alzheimer’s disease. Clin Proteomics 17, 19.

[77] Konijnenberg E, Tijms BM, Gobom J, Dobricic V, Bos I, Vos S, Tsolaki M, Verhey F, Popp J, Martinez-Lage P, Vandenberghe R, Lleo A, Frolich L, Lovestone S, Streffer J, Bertram L, Blennow K, Teunissen CE, Veerhuis R, Smit AB, Scheltens P, Zetterberg H, Visser PJ (2020) APOE epsilon4 genotype-dependent cerebrospinal fluid proteomic signatures in Alzheimer’s disease. Alzheimers Res Ther 12, 65.

[78] Reus LM, Stringer S, Posthuma D, Teunissen CE, Scheltens P, Pijnenburg YAL, Visser PJ, Tijms BM, Alzheimer’s Disease Neuroimaging I (2020) Degree of genetic liability for Alzheimer’s disease associated with specific proteomic profiles in cerebrospinal fluid. Neurobiol Aging 93, 144 e141–144 e115.

[79] Lin YT, Seo J, Gao F, Feldman HM, Wen HL, Penney J, Cam HP, Gjoneska E, Raja WK, Cheng J, Rueda R, Kritskiy O, Abdurrob F, Peng Z, Milo B, Yu CJ, Elmsaouri S, Dey D, Ko T, Yankner BA, Tsai LH (2018) APOE4 Causes Widespread Molecular and Cellular Alterations Associated with Alzheimer’s Disease Phenotypes in Human iPSC-Derived Brain Cell Types. Neuron 98, 1141–1154 e1147.

[80] Vignesh P, Rawat A, Sharma M, Singh S (2017) Complement in autoimmune diseases. Clin Chim Acta 465, 123–130.

[81] Yin C, Ackermann S, Ma Z, Mohanta SK, Zhang C, Li Y, Nietzsche S, Westermann M, Peng L, Hu D, Bontha SV, Srikakulapu P, Beer M, Megens RTA, Steffens S, Hildner M, Halder LD, Eckstein HH, Pelisek J, Herms J, Roeber S, Arzberger T, Borodovsky A, Habenicht L, Binder CJ, Weber C, Zipfel PF, Skerka C, Habenicht AJR (2019) ApoE attenuates unresolvable inflammation by complex formation with activated C1q. Nat Med 25, 496–506.

[82] Hong S, Beja-Glasser VF, Nfonoyim BM, Frouin A, Li S, Ramakrishnan S, Merry KM, Shi Q, Rosenthal A, Barres BA, Lemere CA, Selkoe DJ, Stevens B (2016) Complement and microglia mediate early synapse loss in Alzheimer mouse models. Science 352, 712–716.

[83] Konijnenberg E, Tijms BM, Gobom J, Dobricic V, Bos I, Vos S, Tsolaki M, Verhey F, Popp J, Martinez-Lage P, Vandenberghe R, Lleó A, Frölich L, Lovestone S, Streffer J, Bertram L, Blennow K, Teunissen CE, Veerhuis R, Smit AB, Scheltens P, Zetterberg H, Visser PJ (2020) APOE ε4 genotype-dependent cerebrospinal fluid proteomic signatures in Alzheimer’s disease. Alzheimers Res Ther 12, 65.

[84] Chung WS, Verghese PB, Chakraborty C, Joung J, Hyman BT, Ulrich JD, Holtzman DM, Barres BA (2016) Novel allele-dependent role for APOE in controlling the rate of synapse pruning by astrocytes. Proc Natl Acad Sci U S A 113, 10186–10191.

[85] Mayilyan KR (2012) Complement genetics, deficiencies, and disease associations. Protein Cell 3, 487–496.

